# Elevated glycoprotein acetyl levels in adolescence and early adulthood predict adverse cardiometabolic profiles and risk of metabolic syndrome in up to 10 year follow-up

**DOI:** 10.1101/2020.09.30.20204479

**Authors:** Scott T. Chiesa, Marietta Charakida, Georgios Georgiopoulos, Justin D. Roberts, Simon J. Stafford, Chloe Park, Juha Mykkänen, Mika Kähönen, Terho Lehtimäki, Mika Ala-Korpela, Olli Raitakari, Alun D. Hughes, Naveed Sattar, Nicholas J. Timpson, John E. Deanfield

**Author notes:** **Address for Correspondence:** Scott T Chiesa, PhD, Institute of Cardiovascular Science, University College London, 1 St Martin’s Le Grand, London, UK, EC1A 4NP.

## Abstract

**Objective:** Low-grade inflammation in the young may contribute to the early development of adverse cardiometabolic risk profiles. We assessed whether measures of glycoprotein acetylation (GlycA) were better able to detect the development of these changes compared to the more commonly used biomarker high-sensitivity C-reactive protein (CRP), and investigated whether these relationships differed in an adolescent compared to young adult cohort.

**Research Design and Methods:** A total of 3306 adolescents (Avon Longitudinal Study of Parents and Children - ALSPAC; mean age 15.4±0.3; n=1750) and young adults (Cardiovascular Risk in Young Finns Study - YFS; mean age 32.1±5.0; n=1556) were included. Inflammatory biomarkers (GlycA/CRP), body composition (BMI / waist circumference) and cardiometabolic risk factors (blood pressure, triglycerides, HDL-c, glucose, insulin, and homeostasis model of insulin resistance [HOMA_IR]), were measured at baseline and again in 9-10 year follow-up. Metabolic Syndrome (MetS) was defined using adolescent-specific National Cholesterol Education Programme (NCEP) guidelines in ALSPAC and standard NCEP guidelines in YFS.

**Results:** GlycA levels showed greater within-subject correlation over the 9-10 year duration of follow-up in both cohorts when compared to CRP, particularly in the younger adolescent group. In adjusted models, only GlycA was found to increase in line with cardiometabolic risk factor burden at baseline, and to predict adverse changes in several cardiometabolic risk factors in follow-up. In both cohorts, GlycA predicted future risk of MetS (OR [95%CI] for Q4 vs. Q1 = 1.95 [1.08,3.53] and 2.74 [1.30,5.73] for ALSPAC and YFS, respectively), whereas CRP showed a neutral or even negative relationship in fully-adjusted models (OR [95%CI] = 0.50 [0.29,0.86] and 0.93 [0.53,1.64]).

**Conclusions:** Chronic inflammation is associated with adverse cardiometabolic risk profiles from as early as adolescence and predicts risk of future cardiometabolic risk and MetS in up to 10 year follow-up. GlycA may be a more sensitive inflammatory biomarker to CRP for detecting early cardiometabolic and cardiovascular risk in the young.

## INTRODUCTION

Chronic low-grade inflammation is a hallmark of both cardiometabolic and cardiovascular disease, and is most commonly quantified in clinical research using high-sensitivity assays of C-reactive protein (CRP). Although likely not a causal risk factor itself(1), CRP lies downstream of multiple inflammatory pathways implicated in chronic disease, and has been shown in older populations to predict risk of future incidence of type 2 diabetes and cardiovascular disease(2,3). Whether low-grade inflammation contributes to the emergence of the earliest signs of cardiometabolic dysfunction in the young remains unclear, however, largely due to a lack of randomized clinical trials and/or Mendelian randomization studies at this age.

In cross-sectional analyses, some – but not all – studies have reported modest associations between CRP and various early signs of cardiometabolic disease(4–6). These relationships often disappear following adjustment for potential confounders such as BMI or waist circumference, however, suggesting either one of two things – 1) that inflammation predominantly contributes to the cardiometabolic/cardiovascular disease process in the later stages of disease, and early associations may simply result from confounding due to obesity, or 2) that different inflammatory pathways to those upstream of CRP may underlie early cardiometabolic changes in the young, and novel biomarkers may be required for monitoring low-grade inflammatory burden at this age.

Recent research has identified a potential role for a nuclear magnetic resonance (NMR)-derived measure of glycoprotein acetylation – termed GlycA – as a novel biomarker of systemic inflammation(7). The GlycA NMR signal represents the integrated concentration and glycosylation of numerous acute phase proteins (predominantly alpha-1-acid glycoprotein, haptoglobin, and alpha-1-antitrypsin) released in the inflammatory state. This novel biomarker has recently been shown in multiple older cohorts to predict future development of type 2 diabetes and cardiovascular disease independently of CRP(8,9), suggesting that it may capture different upstream inflammatory pathways relevant to disease. No study to-date, however, has assessed the ability of these biomarkers to predict the early development of well-established cardiometabolic and cardiovascular risk factors such as obesity, hypertension, dyslipidemia, and metabolic dysfunction in a young population. These adverse cardiometabolic changes frequently emerge during the transition from adolescence to young adulthood – both in isolation and clustered together in a condition known as the Metabolic Syndrome (MetS). With long-term exposure to cumulative risk factor burden increasingly being recognized as a major contributor to later-life chronic disease(10), and up to 20% of young adults now recognized to have a clustering of risk factors defined as MetS(11), the identification of factors driving the appearance of cardiometabolic dysfunction in the early stages of life is becoming increasingly important.

Utilising two large and extensively-phenotyped longitudinal cohorts of adolescents and young adults, we now report the first findings comparing relationships between inflammatory biomarkers and cardiometabolic risk factors during the transition from adolescence to mid-life, and investigate the ability of each biomarker to predict the risk of adverse cardiometabolic risk profiles and MetS in up to 10 year follow-up.

## METHODS

### Study Population

Participants were drawn from two ongoing longitudinal cohorts of young people based in the UK and Finland (Avon Longitudinal Study of Parents and Children – ALSPAC; Cardiovascular Risk in Young Finns Study – YFS). ALSPAC is a prospective birth cohort study investigating factors that influence normal childhood development and growth, whereas YFS is a multi-center follow-up study conducted in five Finnish cities (Helsinki, Kuopio, Oulu, Tampere, and Turku) and their rural surroundings. Cohort and study designs for both studies have been described in detail previously(12–15), and a brief description of each is provided in the Supplemental File. In ALSPAC, ethical approval was obtained from the ALSPAC Ethics and Law Committee and Local Research Ethics Committees. If the child was younger than age 16 years at the time of consent, informed written consent was obtained from the parent/guardian alongside assent from the child. When age 16 years or older, all participants provided their own informed written consent. In YFS, written informed consent was obtained from local research ethics committees. All ethical approvals from both cohorts conformed to the Declaration of Helsinki and all biological samples used in the study were collected in accordance with the Human Tissue Act (2004).

### Study Design

The ability of inflammatory biomarkers in adolescence to predict adverse cardiometabolic profiles and MetS in young adulthood was tested in ALSPAC using data collected from individuals who attended both Teen Focus 3 (mean age 15 years) and Focus at 24 (mean age 24 years) clinics. Data collected from YFS participants attending both 2001 (mean age 32 years) and 2011 (mean age 42 years) follow-up clinics were subsequently used to test the same hypotheses during the transition from early- to mid-adulthood. Only participants with both inflammatory markers measured at baseline and cardiometabolic risk factors at follow-up were included. Any participants with CRP levels > 10mg/L were excluded in order to reduce the risk of confounding arising due to acute infection, leaving a total of 3307 individuals (ALSPAC n=1751 and YFS n = 1556) in the study.

### Inflammatory and Cardiometabolic Risk Factors

GlycA in both studies was measured as part of an NMR metabolomics platform (Nightingale Health, Helsinki, Finland). High-sensitivity CRP was measured by automated particle-enhanced immunoturbidimetric assay in ALSPAC (Roche UK, Welwyn Garden City, UK), and in YFS by an Olympus AU400 with “CRP-UL” assay kit (Wako Chemicals, Neuss, Germany). Body mass index (BMI) was calculated as weight(kg)/height(m)^2^ and waist circumference was measured using a flexible tape to the nearest 1 mm at the midpoint between the lower ribs and the iliac crest. Blood pressure (BP) was measured in the seated position in ALSPAC and supine position in YFS using an automated sphygmomanometer. Blood lipids, glucose, and insulin were all measured as previously described(16), and insulin resistance was estimated using the Homeostasis Model Assessment (HOMA2-IR, Diabetes Trials Unit, Oxford).

### Definition of Metabolic Syndrome (MetS)

MetS in ALSPAC was characterised using a modified version of the National Cholesterol Education Programme (NCEP) guidelines due to the young age of participants and their transition through adolescence during the course of follow-up. In this version, a participant was classified as having MetS if they had any three of the following five age- and sex-specific components: elevated waist circumference ≥80th percentile, systolic or diastolic blood pressure ≥ 80th percentile, triglycerides ≥80th percentile, glucose ≥80th percentile, or HDL-C ≤ 20th percentile. In the older YFS cohort, standard NCEP guidelines were used: waist >102 cm in men and >88 cm in women, serum triglycerides ≥1.695 mmol/l (150 mg/dl), HDL cholesterol <1.036 mmol/l (40 mg/dl) in men and 1.295 mmol/l (50 mg/dl) in women, blood pressure ≥ 130mmHg (systolic) or ≥85mmHg (diastolic) or treated, and plasma glucose ≥5.6 mmol/l (110 mg/dl). To complement the dichotomous definitions, a continuous MetS risk score was also calculated to represent summative cardiometabolic risk factor burden. For the continuous score, the components were first standardized (z-scored) for age and sex (HDL-C was multiplied by −1) and then summed by age. As a representation of blood pressure, the mean of the systolic and diastolic blood pressure values was used.

### Lifestyle Risk Factors

In ALSPAC, highest household occupation was used to assign participants a household social class, using the 1991 British Office of Population Census Statistics classification(17), while physical activity was defined as the average counts per minute (CPM) recorded over 7 days using an MTI Actigraph AM7164 2.2 accelerometer in a subsample of participants. In YFS, socioeconomic position was assessed using current occupational status (manual/ lower-grade non-manual / higher-grade non-manual), and physical activity using a physical activity index (PAI) generated from questionnaire data detailing exercise habits and frequency, as previously described(18).

### Statistical Analysis

Continuous data were summarized as mean ± SD or median (interquartile range) if skewed. Normal distribution was assessed using the Shapiro–Wilk test alongside graphical inspection of histograms and normality plots, and non-normally distributed data were log-transformed prior to inclusion in statistical models. In cross-sectional analyses, Pearson correlation was first used to assess bivariate relationships between inflammatory biomarkers and cardiometabolic risk factors at baseline. Additional relationships between adverse cardiometabolic profiles and inflammatory biomarker levels were then also assessed using multivariable linear regression models, first unadjusted and then adjusted for age, sex, and BMI. In this analysis, the number of MetS risk factors present at baseline was used as the independent variable, while the mean increase and 95% confidence intervals (CIs) in each biomarker compared to individuals with zero components (reference group) was used as the outcome. In longitudinal analyses, bivariate scatter plots were used to graphically represent correlations between GlycA and CRP levels measured on two separate occasions up to 10 years apart using both absolute and log-transformed values. The ability for high and low levels of inflammatory biomarkers at baseline (lower quartile vs. upper quartile [Q1 v Q4]) to predict differences in cardiometabolic risk factors at follow-up were then assessed using multivariable linear regression analysis adjusted for baseline age, sex, BMI, waist circumference, HDL-c, triglycerides, glucose, blood pressure, other inflammatory marker, baseline physical activity levels and socioeconomic status. All outcomes were reported as z-scores to aid comparison between different risk factors. Next, logistic regression analyses using baseline values of GlycA or CRP as exposures were used to calculate odds of a diagnosis of MetS at study follow-up. Both continuous (per z-score change) and threshold (quartiles) relationships were tested, and results were expressed as odds ratios (OR) and 95% confidence intervals. Four models were created for each inflammatory biomarker: Model 1 = unadjusted; Model 2 = model 1 + adjustments for baseline age, sex, and BMI; Model 3 = model 2 + adjustments for baseline waist circumference, HDL-c, triglycerides, glucose, blood pressure, and other inflammatory marker; and Model 4 = model 3 + adjustments for baseline physical activity levels and socioeconomic status. Similar models excluding participants already classed as having MetS at baseline were used to test for incidence of new diagnoses during the course of the study. To reduce risk of confounding by acute infection, participants self-reporting acute illness in the previous 3 weeks or with CRP levels > 10 mg/dl were removed from analyses (n=52 for ALSPAC and n=89 for YFS). Retention of these participants, however, had no noticeable effect on results (data not shown). In all analyses, multiple imputations (10 imputed datasets) were used to account for missing covariates in statistical models. Details of missing data in both cohorts are shown in Supplementary Table 1. Tests for interactions between sex and inflammatory biomarkers were non-significant (p > 0.25), and all results are therefore presented with males and females combined. All statistical analyses were conducted using Stata 15.1 (StataCorp LLC, Texas, USA). A priori, we planned to draw conclusions based on effect estimates and their 95% CIs, rather than statistical tests using an arbitrary p-value cutoff, although these are still provided for reference. For example, given two effects with the same point estimate— one with narrow CIs, the other with wider CIs that may even include the null—we described both as showing the same effect. However, one is more imprecisely estimated and should be treated with more caution until replicated in a larger sample.

## RESULTS

### Participant Characteristics

Participants in ALSPAC were mean age of 15.4 ± 0.3 years at baseline and 57% female, whereas those in YFS were on average 32.1 ± 5.0 years of age and 55% female. Prevalence of MetS increased progressively with age, from 6.9% at age 15 to 18% at age 42 (Table 1). All other characteristics are shown in Table 1.

**Table 1:** Participant characteristics in adolescent and young adult cohorts. ALSPAC = Avon Longitudinal Study of Parents and Children; YFS = Cardiovascular Risk in Young Finns Study; BMI = body mass index; SBP = systolic blood pressure; DBP = diastolic blood pressure; MAP = mean arterial pressure; LDL-c = low-density lipoprotein cholesterol; HDL-c = high-density lipoprotein cholesterol; HOMA2-IR = homeostatic model of assessment for insulin resistance; GlycA = glycoprotein acetyls; CRP = high-sensitivity C-reactive protein; MetS = Metabolic Syndrome; CPM = counts per minute; PAI = physical activity index

| Variable | ALSPAC |  |  | YFS |  |  |
| --- | --- | --- | --- | --- | --- | --- |
|  | n | Baseline | Follow Up | n | Baseline | Follow Up |
| Age (years) | 1748 | 15.4 ± 0.3 | 24.0 ± 0.8 | 1556 | 32.1 ± 5.0 | 42.1 ± 5.0 |
| Sex (% female) | 1750 | 57.2 | 57.2 | 1556 | 55.0 | 55.0 |
| Height (m) | 1735 | 1.69 ± 0.08 | 1.72 ± 0.09 | 1547 | 1.72 ± 0.9 | 1.72 ± 0.9 |
| Mass (kg) | 1750 | 60.6 ± 13.2 | 73.3 ± 17.9 | 1547 | 74.5 ± 16.0 | 78.9 ± 17.3 |
| BMI (kg/m<sup>2</sup>) | 1735 | 20.6 (18.9, 22.7) | 23.6 (21.4, 26.8) | 1547 | 24.3 (22.0, 27.3) | 25.7 (23.1, 29.1) |
| Waist Circumference (cm) | 1462 | 76.3 ± 8.5 | 80.6 ± 14.9 | 1530 | 84.0 ± 12.3 | 91.7 ± 14.1 |
| SBP (mmHg) | 1691 | 123 ± 11 | 116 ± 13 | 1540 | 122 ± 14 | 126 ± 15 |
| DBP (mmHg) | 1691 | 66 ± 10 | 67 ± 8 | 1540 | 73 ± 9 | 79 ± 11 |
| MAP (mmHg) | 1691 | 85 ± 8 | 83 ± 8 | 1540 | 89 ± 10 | 94 ± 12 |
| LDL-c (mmol/l) | 1750 | 2.1 ± 0.6 | 2.5 ± 0.8 | 1538 | 3.3 ± 0.8 | 3.3 ± 0.8 |
| HDL-c (mmol/l) | 1750 | 1.3 ± 0.3 | 1.6 ± 0.4 | 1555 | 1.3 ± 0.3 | 1.3 ± 0.3 |
| Triglycerides (mmol/l) | 1750 | 0.8 (0.6, 1.0) | 0.8 (0.7, 1.2) | 1556 | 1.1 (0.8, 1.6) | 1.1 (0.8, 1.6) |
| Glucose (mmol/l) | 1750 | 5.2 ± 0.4 | 5.3 ± 0.7 | 1556 | 5.0 ± 0.8 | 5.4 ± 1.0 |
| Insulin (µU/ml) | 1750 | 9.0 (6.7, 11.8) | 7.5 (5.3, 10.9) | 1556 | 6.0 (5.0, 9.0) | 7.3 (4.4, 11.5) |
| HOMA2-IR | 1750 | 1.2 (0.9, 1.5) | 1.0 (0.7, 1.4) | 1556 | 0.8 (0.6, 1.2) | 1.0 (0.6, 1.5) |
| GlycA (mmol/L) | 1750 | 1.19 (1.12, 1.27) | 1.21 (1.11, 1.33) | 1556 | 1.34 (1.21, 1.52) | 1.56 (1.42, 1.73) |
| CRP (mg/l) | 1750 | 0.34 (0.21, 0.80) | 0.85 (0.39, 2.26) | 1556 | 0.71 (0.31, 1.67) | 0.75 (0.34, 1.66) |
| MetS (%) | 1750 | 6.9 | 11.2 | 1556 | 10.8 | 18.0 |
| Physical Activity<br>(CPM)<br>(PAI) | 922<br>- | 472 ± 170<br>- | -<br>- | -<br>1556 | -<br>8.86 ± 1.97 | -<br>- |
| Highest Household Social Class (%) | 1456 |  |  | 1522 |  |  |
| - I |  | 9.1 | - |  | - | - |
| - II |  | 37.8 | - |  | - | - |
| - III (non-manual) |  | 40.4 | - |  | - | - |
| - III (manual) |  | 5.4 | - |  | - | - |
| - IV |  | 6.0 | - |  | - | - |
| - V |  | 1.3 | - |  | - | - |
| - Higher Grade Non-Manual |  | - | - |  | 27.0 | - |
| - Lower Grade Non-Manual |  | - | - |  | 43.2 | - |
| - Manual |  | - | - |  | 29.8 | - |

### Cross-Sectional Relationships Between Inflammatory Biomarkers and Cardiometabolic Risk Factors at Baseline

GlycA in both cohorts was most strongly associated with body composition, triglycerides, insulin, and HOMA-IR. CRP in the younger ALSPAC cohort showed little relationship to any risk factor with the exception of body composition, whereas in the older YFS cohort it also demonstrated moderate relationships with insulin and HOMA-IR (Figure 1). GlycA levels in both cohorts increased in line with an increasing number of cardiometabolic risk factors, with this relationship remaining essentially unchanged following adjustment for age, sex, and BMI. In contrast, any observed increase in CRP levels in both cohorts were attenuated towards the null after accounting for differences in BMI between groups (Figure 2).

**Figure 1:**
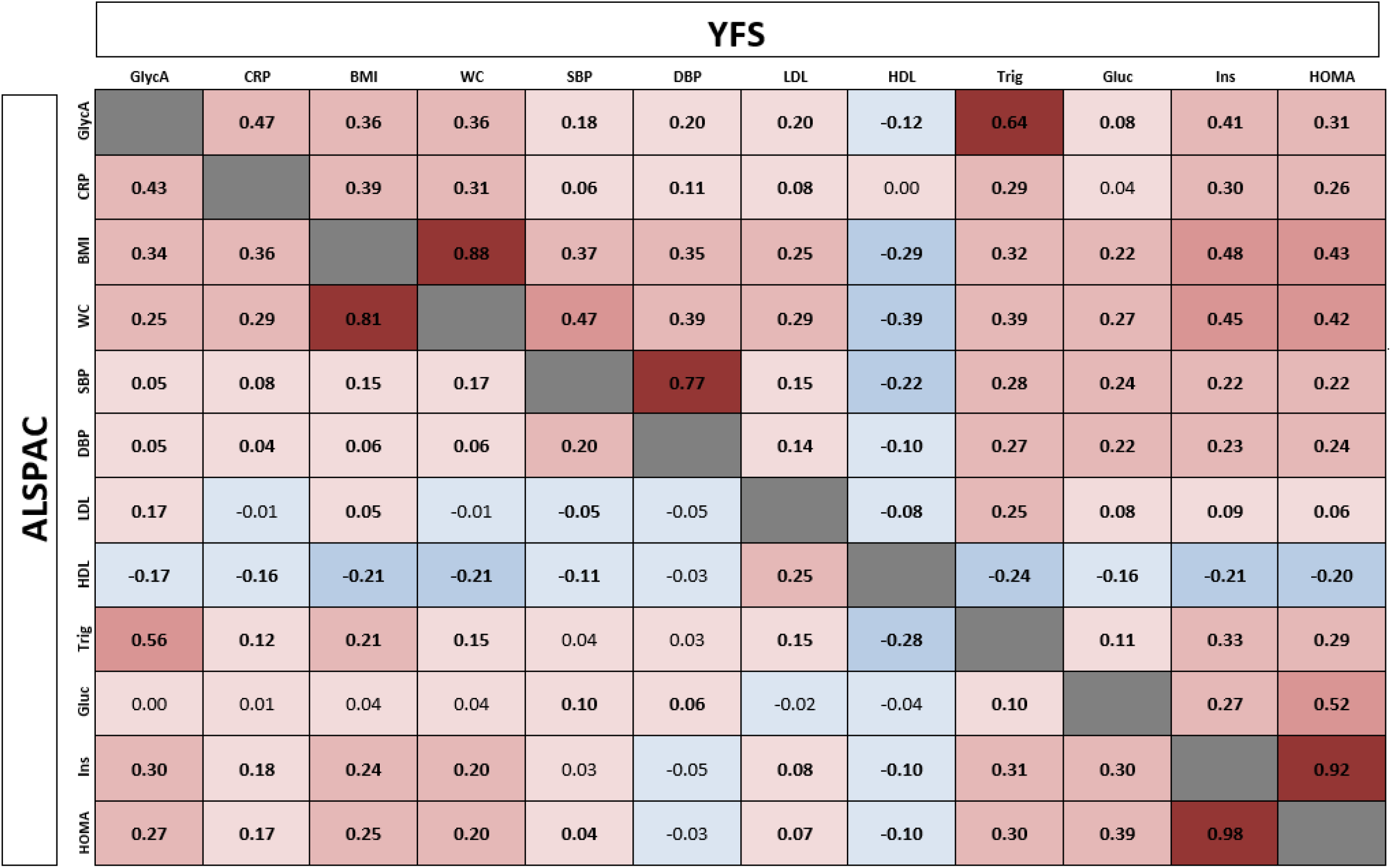
Cross-sectional relationships between inflammatory biomarkers and cardiometabolic risk factors in ALSPAC and YFS cohorts. Bold text indicates p < 0.05. GlycA, CRP, triglycerides, and insulin all log-transformed prior to inclusion.

**Figure 2:**
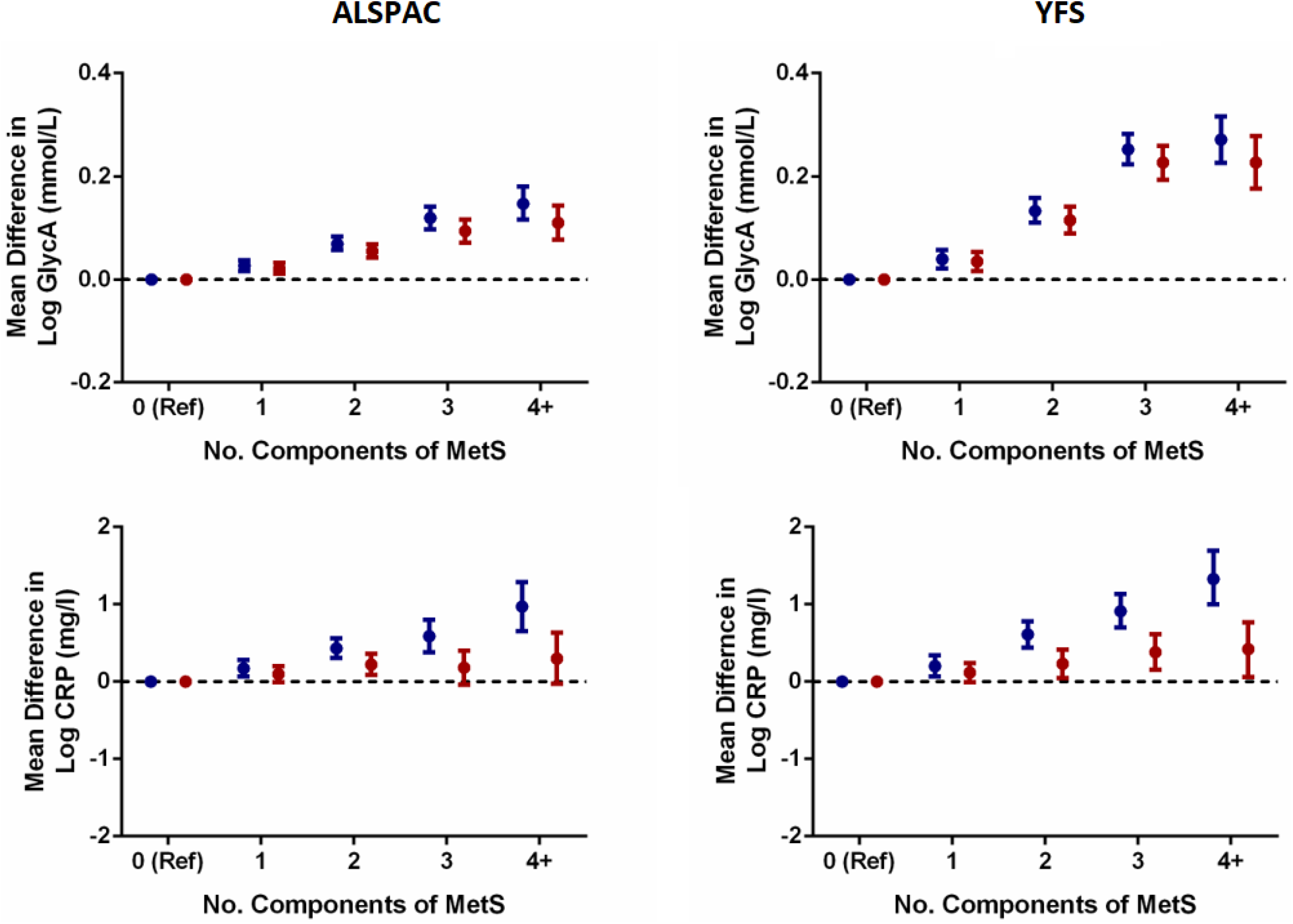
Cross-sectional relationships between number of components of MetS and inflammatory biomarker levels in ALSPAC and YFS cohorts. Plots represent mean difference from reference group (zero MetS components) and 95% confidence intervals. Blue = unadjusted; Red = adjusted for age, sex, and BMI

### Inflammatory Biomarker Levels in Acute Illness and Stability Over Time

Levels of inflammatory biomarkers tended to be lower in adolescence compared to young adulthood (Table 1). Individuals reporting an acute infectious illness in the previous three weeks in the ALSPAC cohort had increased levels of both biomarkers compared to those without, with this effect particularly pronounced for CRP (5% vs. 121% increase for GlycA and CRP, respectively; p < 0.001 for both; Supplemental Figure 1). Similar findings have previously been reported for the YFS cohort(19). After excluding these individuals, repeat measures of GlycA taken 9-10 years apart showed a moderate correlation in both cohorts (r = 0.41 and 0.36 for ALSPAC and YFS, respectively; p < 0.001 for both; Supplemental Figure 2), whereas repeat measures of CRP showed little relationship in ALSPAC (r = 0.07) and only a moderate correlation in YFS (r = 0.30; Supplemental Figure 2). Log transformation of data had little impact on GlycA, but strengthened the relationship between repeat measures of CRP (Supplemental Figure 3).

### Longitudinal Relationships Between Inflammatory Biomarkers at Baseline and Individual Cardiometabolic Risk Factors in 9-10 Year Follow-Up

In the adolescent ALSPAC cohort, elevated GlycA at baseline (Q4 vs. Q1) was an independent predictor of numerous adverse cardiometabolic risk factors at follow-up – in particular higher levels of diastolic blood pressure, glucose, and insulin resistance – whereas elevated levels of CRP showed evidence of reduced levels of these same risk factors (Figure 3). In the older YFS cohort, GlycA showed similar associations with future elevations in DBP alongside some evidence of dyslipidemia, whereas no relationship was observed between CRP and any cardiometabolic risk factor. Regression models conducted without using imputed values for physical activity and socioeconomic class (covariates with high levels of missingness) gave near identical results.

**Figure 3:**
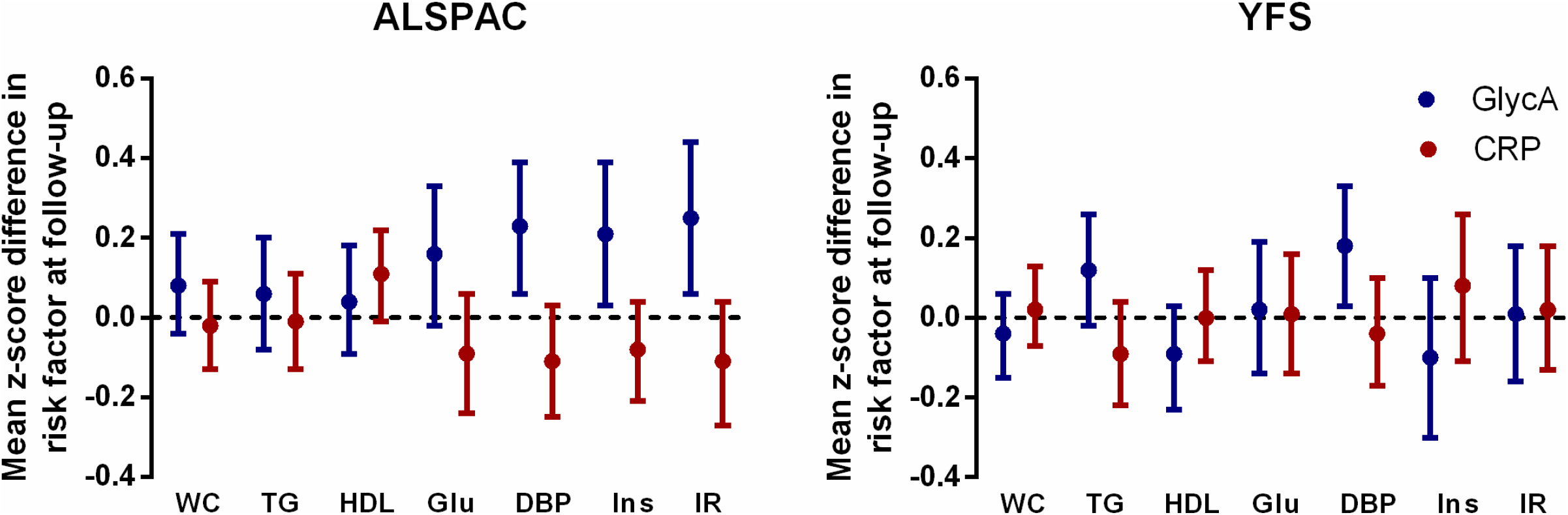
Predictive value of high and low levels of inflammatory biomarkers for cardiometabolic risk factors measured 9-10 years later. Plots represent mean difference and 95% confidence intervals for each risk factor at follow-up when comparing highest (Q4) vs. lowest (Q1) levels of each inflammatory biomarker at baseline. Relationships between each biomarker and risk factor adjusted for age, sex, BMI, each of the other risk factors and biomarkers, physical activity, and socioeconomic status. WC – waist circumference; TG – triglycerides; Glu – glucose; DBP – diastolic blood pressure; Ins – insulin; IR – HOMA insulin resistance

### Predictive Value of Inflammatory Biomarkers for Detecting Risk of MetS in 9-10 Year Follow-Up

In unadjusted models, both biomarkers were associated with an increased risk of future MetS in both linear and threshold analyses (Table 2). These relationships were stronger for GlycA than CRP and were particularly pronounced in the older YFS cohort (OR [95%CI] = 4.25 [2.73, 6.61] vs. 2.18 [1.44, 3.28] for Q4 vs. Q1 for GlycA and CRP respectively in ALSPAC; and 14.79 [8.34, 26.21] vs. 3.72 [2.48, 5.57] for the same measures in YFS). In fully adjusted models, only GlycA predicted future risk of MetS (OR [95%CI] for Q4 vs. Q1 = 1.95 [1.08, 3.53] and 2.74 [1.30, 5.73] in ALSPAC and YFS, respectively). CRP showed a negative relationship to MetS in ALSPAC (OR [95%CI] = 0.50 [0.29, 0.86]) and a neutral relationship in YFS (OR = 0.93 [0.53, 1.64]; Table 2). Consistent results were observed when using a summative risk score for MetS rather than dichotomous values (Supplemental Table 2) and when excluding participants with prevalent MetS at baseline (Supplemental Table 3). Regression models conducted without using imputed values for physical activity and socioeconomic class again gave near identical results.

**Table 2:** Predictive value of inflammatory biomarkers for detecting risk of MetS in 9-10 year follow-up. Model 1 = unadjusted; Model 2 = model 1 + adjustments for baseline age, sex, and BMI; Model 3 = model 2 + adjustments for baseline waist circumference, HDL, triglycerides, glucose, blood pressure, and other inflammatory marker; Model 4 = model 3 + adjustments for baseline physical activity levels and socioeconomic status. * p < 0.05 ^#^ p < 0.01 † p < 0.001

| ALSPAC |  |  |  |  |  |  |
| --- | --- | --- | --- | --- | --- | --- |
|  | Quartile 1 | Quartile 2 | Quartile 3 | Quartile 4 |  | Per 1 SD |
| ***GlycA*** |  |  |  |  |  |  |
| Model 1 | 1 (Ref) | 1.45 (0.88, 2.39) | 1.64 (1.01, 2.67)* | 4.25 (2.73, 6.61)† |  | 1.70 (1.49, 1.95)† |
| Model 2 | 1 (Ref) | 1.26 (0.76, 2.09) | 1.15 (0.69, 1.92) | 2.28 (1.40, 3.75)<sup>#</sup> |  | 1.35 (1.15, 1.57)† |
| Model 3 | 1 (Ref) | 1.14 (0.67, 1.92) | 1.08 (0.62, 1.86) | 2.00 (1.11, 3.60)* |  | 1.31 (1.05, 1.63)† |
| Model 4 | 1 (Ref) | 1.13 (0.67, 1.91) | 1.05 (0.60, 1.82) | 1.95 (1.08, 3.53)* |  | 1.33 (1.06, 1.66)† |
| ***CRP*** |  |  |  |  |  |  |
| Model 1 | 1 (Ref) | 1.15 (0.74, 1.79) | 1.70 (1.12, 2.59)* | 2.18 (1.44, 3.28)† |  | 1.05 (0.93, 1.18) |
| Model 2 | 1 (Ref) | 0.84 (0.53, 1.33) | 0.98 (0.62, 1.53) | 0.76 (0.47, 1.24) |  | 0.92 (0.76, 1.11) |
| Model 3 | 1 (Ref) | 0.82 (0.51, 1.33) | 0.82 (0.51, 1.33) | 0.51 (0.30, 0.87)* |  | 0.78 (0.59, 1.02) |
| Model 4 | 1 (Ref) | 0.81 (0.50, 1.32) | 0.82 (0.51, 1.33) | 0.50 (0.29, 0.86)* |  | 0.76 (0.57, 1.01) |
| YFS |  |  |  |  |  |  |
|  | Quartile 1 | Quartile 2 | Quartile 3 | Quartile 4 |  | Per 1 SD |
| ***GlycA*** |  |  |  |  |  |  |
| Model 1 | 1 (Ref) | 3.42 (1.84, 6.34)† | 7.24 (4.03, 13.01)† | 14.79 (8.34, 26.21)† |  | 2.14 (1.88, 2.44)† |
| Model 2 | 1 (Ref) | 2.54 (1.34, 4.83)<sup>#</sup> | 4.69 (2.55, 8.64)† | 7.34 (4.01, 13.42)† |  | 1.84 (1.58, 2.13)† |
| Model 3 | 1 (Ref) | 2.05 (1.04, 4.06)* | 2.92 (1.49, 5.74)<sup>#</sup> | 2.76 (1.32, 5.77)<sup>#</sup> |  | 1.34 (1.08, 1.67)<sup>#</sup> |
| Model 4 | 1 (Ref) | 2.26 (1.13, 4.49)* | 3.08 (1.56, 6.10)<sup>#</sup> | 2.74 (1.30, 5.73)<sup>#</sup> |  | 1.33 (1.07, 1.67)* |
| ***CRP*** |  |  |  |  |  |  |
| Model 1 | 1 (Ref) | 1.70 (1.10, 2.64)* | 2.46 (1.62, 3.74)† | 3.72 (2.48, 5.57)† |  | 1.22 (1.09, 1.37)† |
| Model 2 | 1 (Ref) | 1.39 (0.88, 2.20) | 1.46 (0.93, 2.31) | 1.57 (0.97, 2.53) |  | 1.02 (0.88, 1.18) |
| Model 3 | 1 (Ref) | 1.08 (0.64, 1.82) | 0.97 (0.57, 1.64) | 0.88 (0.50, 1.55) |  | 0.93 (0.78, 1.11) |
| Model 4 | 1 (Ref) | 1.12 (0.66, 1.89) | 0.98 (0.58, 1.67) | 0.93 (0.53, 1.64) |  | 0.92 (0.76, 1.09) |

## DISCUSSION

In two extensively-phenotyped longitudinal cohorts containing over 3300 adolescents and young adults with repeat measures of inflammatory and cardiometabolic risk factors over 9-10 years of follow-up, we provide the first evidence of a relationship between early chronic low-grade inflammation and long-term risk of adverse cardiometabolic profiles and MetS. Our findings show that 1) GlycA is a stable marker of chronic inflammation over time and is less sensitive to acute infection compared to CRP, particularly in adolescence; 2) unlike CRP, associations between GlycA and cardiometabolic risk factors are not confounded by BMI; and 3) GlycA is a superior biomarker to CRP for predicting long-term development of adverse cardiometabolic profiles and MetS at this early age, with this association once again particularly evident during adolescence.

Inflammation is increasingly recognized as a hallmark for both type 2 diabetes and cardiovascular disease(20), and increasing evidence suggests that life-time rather than contemporary exposure likely poses the greatest risk for future disease prevalence and events (21). Despite a long-hypothesized link between chronic inflammation and the early development of adverse cardiometabolic risk profiles, studies using the inflammatory biomarker CRP to link these factors have to-date produced equivocal results(4,22,23). In the current study, we therefore sought to compare and contrast prospective associations between CRP and a novel inflammatory biomarker (GlycA) for the prediction of future cardiometabolic risk in two independent cohorts spanning an age-range from adolescence to mid-adulthood. Our findings support a number of lines of prior evidence suggesting that GlycA may be a more suitable measure of early chronic inflammatory burden in early life. Firstly, GlycA has previously been shown to be considerably more stable over time than CRP(19), with the latter often virtually undetectable in young healthy individuals except in the case of acute infections(24). These findings were confirmed in the current study, where repeat measures of GlycA taken up to 10 years apart showed stronger correlations than those seen for CRP. This was particularly evident in the younger adolescent cohort, where overall CRP levels were very low, with < 5% of individuals found to have levels above the 3mg/L level commonly used as a threshold for increased risk in older or clinical populations(25). As well as being more stable over time, GlycA was also found to be less sensitive to acute infection, increasing on average by only ∼5% in individuals reporting acute illness in the previous 3 weeks, compared to 120% for CRP. Taken together, these findings suggest that GlycA and CRP may capture inflammatory burden in a similar manner as that seen for HbA1c and fasting glucose when assessing blood sugar control, with the former more representative of longer-term exposure to cumulative inflammatory burden, and the latter more sensitive to acute changes which accompany infectious illness. Secondly, in the absence of long-term exposure to cardiometabolic risk factors and co-morbidities at an early age, modifiable lifestyle factors such as obesity, physical fitness, and diet are likely to be particularly important potential sources for chronic inflammation in the young. Recent research has shown strong cross-sectional relationships between GlycA and a number of these factors even in childhood, with individuals as young as 11 years old demonstrating associations between GlycA and obesity, physical fitness, diet, and insulin resistance(26,27). In the current study, cross-sectional analyses in both cohorts at ages 15 (ALSPAC) and 32 (YFS) confirmed many of these prior associations, with GlycA found to most closely relate to body composition, triglyceride levels, and markers of insulin resistance; whereas CRP was predominantly related to body composition alone. Similar to previous research, we therefore found any association between CRP and increasing cardiometabolic risk factor burden to be attenuated towards the null after accounting for BMI, suggesting that obesity, rather than inflammatory burden captured by CRP, may be the main driver of cardiometabolic risk in these individuals(4,22). In contrast, however, adjustment for BMI in both cohorts had little impact on GlycA levels, which progressively increased in line with the number of cardiometabolic risk factors present. Together, these findings suggest a potential relationship between separate chronic inflammatory pathways and the emergence of adverse cardiometabolic profiles which may be independent of obesity *per se*, and which remain undetected using traditional inflammatory biomarkers such as CRP.

The question of whether this chronic inflammation occurs as a cause or consequence of elevated cardiometabolic risk, however, remains unresolved. In support of the former, recent genetic analyses from the ALSPAC cohort in individuals with a polygenic risk score predisposing to type 2 diabetes have demonstrated early elevations in GlycA prior to the appearance of numerous other cardiometabolic risk factors, suggesting that the low-grade inflammation captured using this biomarker may precede the appearance of later overt metabolic changes which often emerge during the transition through adolescence(28). In an attempt to investigate this further in the current study, we used both ALSPAC and YFS cohorts to test prospective relationships between inflammatory biomarker levels at baseline and cardiometabolic risk factors 9-10 years later, and found that these relationships differed somewhat between the younger and older cohorts. In ALSPAC, elevations in GlycA were found to predominantly predict hyperinsulinemia and insulin resistance in follow-up, whereas in YFS they were more closely related to dyslipidemia. Despite these differences, however, individuals in the top quartile for GlycA in both cohorts were found to have an equivalent and clinically-meaningful (∼2mmHg) increase in future DBP, suggesting a potential relationship between inflammatory burden captured by GlycA and early vascular dysfunction. In support of this hypothesis, GlycA has previously been shown to predict vascular inflammation over-and-above traditional risk factors in healthy middle-aged individuals(29), although whether this also applies at younger ages requires further testing. Regardless of the differences in underlying risk factors observed between cohorts, elevated levels of GlycA were found to predict future risk of MetS to a similar extent in both cohorts, with a 2-3-fold increased risk found when comparing the highest vs. lowest quartile of GlycA. In contrast, any observed positive associations between CRP and cardiometabolic risk (either as individual risk factors or MetS) were once again lost following adjustment for BMI. Unexpectedly, we found CRP in ALSPAC to in fact be negatively associated with a number of risk factors in fully-adjusted models, and was therefore associated with a decreased risk of MetS during follow-up. The reasons for this are unclear, but are similar to previous findings in this same cohort where CRP has shown some evidence of a negative relationship to carotid intima-media thickness – a commonly-used marker of subclinical atherosclerosis(30).

Whether an increased level of GlycA represents a causal risk factor for disease itself or – like CRP – is simply a biomarker increasing in response to other upstream causal pathways, also remains unclear. Given the heterogenous nature of the acute phase proteins which constitute the GlycA NMR signal, this will prove difficult to test, as two individuals with the same GlycA level may have different concentrations and/or glycosylation patterns in underlying proteins. As each of these proteins in isolation are generally considered to possess anti-inflammatory properties, however, it is reasonable to assume that elevated GlycA levels may represent an ongoing attempt at a protective response to underlying chronic inflammatory burden(31), and that interventions targeting pathways upstream are likely offer the best opportunity for risk reduction. These precise pathways have yet to be elucidated, but may differ depending on an individual’s age or underlying inflammatory burden. For example, in a recent study comparing patients with rheumatoid arthritis to healthy controls, distinctly different relationships were observed between GlycA and risk factors depending on the presence or absence of disease. While GlycA in patients with arthritis was found to strongly associate with disease activity and the classic inflammatory markers IL-6 and CRP, associations in individuals without disease were instead similar to that reported here – i.e. predominantly related to body composition and insulin resistance with little-to-no relationship evident for IL-6 or CRP(32). These findings further support the prospect that in young people free from clinical disease, inflammatory pathways separate to the well-established IL-6/CRP axis may be more important for early disease development. While these pathways remain to be determined, recent research into possible determinants of GlycA levels has implicated a role for an ongoing innate immune response dominated by neutrophil-related processes(19); a finding which is particularly interesting given previous findings highlighting the major role which neutrophils play early in the atherogenic process(33).

This study is not without its limitations. Firstly, as with all cohort studies, it is not possible to definitively determine the direction of causality between observed associations, and further interventional or genetic studies are required to answer this question. Secondly, in the absence of hard disease outcomes at this age such as type 2 diabetes or cardiovascular events, we chose to represent increased cardiometabolic risk using MetS as a dichotomous value. The concept of treating MetS this way (i.e. present or absent) rather than recognizing it as a single point on a continuum of cardiometabolic risk is frequently open to debate. For this reason, we also investigated relationships between each inflammatory biomarker and a summative z-score of overall cardiometabolic risk, both of which gave consistent results. Lastly, while we noted that increased levels of GlycA were associated with increased risk of MetS, a threshold for ‘healthy’ levels of GlycA remains unknown. Future studies using both healthy and clinical cohorts will help to answer this question.

## CONCLUSION

In conclusion, we provide the first evidence of an association between chronic inflammation and future development of adverse cardiometabolic profiles and MetS in two separate cohorts spanning an age range from adolescence to mid-adulthood. These relationships were only detectable using GlycA, which may capture different upstream inflammatory pathways underlying cardiometabolic risk at this age. GlycA may therefore be a more sensitive inflammatory biomarker than CRP for detecting early cardiometabolic and cardiovascular risk in the young.

## Data Availability

The data that support the findings of this study are available from the corresponding author upon reasonable request.

## ACKNOWLEDGEMENTS

We are extremely grateful to all the families who took part in this study, the midwives for their help in recruiting them, and the whole ALSPAC and YFS teams, which includes interviewers, computer and laboratory technicians, clerical workers, research scientists, volunteers, managers, receptionists, and nurses.

## AUTHOR CONTRIBUTIONS

STC reviewed the literature, acquired and interpreted data, performed statistical analysis, and drafted the first version of the manuscript. MC and CP acquired and interpreted data. GG interpreted data and performed statistical analysis. JDR and SJS interpreted data. ADH, NJT, and JED have supported data collection in ALSPAC, whereas JM, MK,TL, MAK, and OR have done the same for YFS. NS performed laboratory analyses on blood samples. All authors were involved in critical discussion throughout the data analysis process, critically reviewed the manuscript during the writing process, and approved the final version. STC is the guarantor of this work and, as such, had full access to all the data in the study and takes responsibility for the integrity of the data and the accuracy of the data analysis.

